# Sulfation of *O*-glycans on mucin-type proteins from serous ovarian epithelial tumors

**DOI:** 10.1101/2021.07.05.21259706

**Authors:** Kristina A. Thomsson, Varvara Vitiazeva, Constantina Mateoiu, Chunsheng Jin, Jining Liu, Jan Holgersson, Birgitta Weijdegard, Karin Sundfeldt, Niclas G. Karlsson

**Affiliations:** Department of Medical Biochemistry and Cell Biology, Institute of Biomedicine, Sahlgrenska Academy, University of Gothenburg, Gothenburg, Sweden; Department of Pathology and Cytology, Institute of Biomedicine, Sahlgrenska Academy, University of Gothenburg, SE-41345, Gothenburg, Sweden; Department of Laboratory Medicine, Institute of Biomedicine, Sahlgrenska Academy, University of Gothenburg, SE-41345, Gothenburg, Sweden; Department of Obstetrics and Gynecology, Institute of Clinical Sciences, Sahlgrenska Center for Cancer Research, University of Gothenburg, Gothenburg, Sweden; Department of Life Sciences and Health, Faculty of Health Sciences, Oslo Metropolitan University, Oslo, Norway

## Abstract

Despite that sulfated *O*-linked glycans are abundant on ovarian cancer (OC) glycoproteins, their regulation during cancer development and involvement in cancer pathogenesis remain unexplored. We characterized *O*-glycans carrying sulfation on galactose residues and compared their expression to defined sulfotransferases regulated during OC development. Desialylated sulfated oligosaccharides were released from acidic glycoproteins in the cyst fluid from one patient with a benign serous cyst and one patient with serous OC. Oligosaccharides characterized by LC-MS^n^ were identified as core 1 and core 2 *O*-glycans up to the size of decamers, and with 1-4 sulfates linked to GlcNAc residues and to C-3 and/or C-6 of Gal. To study the specificity of the potential ovarian sulfotransferases involved, Gal3ST2 (Gal-3S)-, Gal3ST4 (Gal-3S)-, and CHST1 (Gal-6S)-encoding expression plasmids were transfected individually into CHO cells also expressing the P-selectin glycoprotein ligand-1/mouse immunoglobulin G2b (PSGL-1/mIg G2b) fusion protein and the human core 2 transferase (GCNT1). Characterization of the PSGL-1/mIg G2b *O*-glycans showed that Gal3ST2 preferentially sulfated Gal on the C-6 branch of core 2 structures and Gal3ST4 preferred Gal on the C-3 branch independently if core-1 or-2. CHST1 sulfated Gal residues on both the C-3 (core 1/2) and C-6 branches of core 2 structures. Using serous ovarian tissue micro array, Gal3ST2 was found to be decreased in tissue classified as malignant compared to tissues classified as benign or borderline, with the lowest expression in poorly differentiated malignant tissue. Neither Gal3ST4 nor CHST1 were differentially expressed in benign, borderline or malignant tissue, and there was no correlation between expression level and differentiation stage. The data displays a complex sulfation pattern of *O*-glycans on OC glycoproteins and that aggressiveness of the cancer is associated with a decreased expression of the Gal3ST2 transferase.

## INTRODUCTION

Epithelial ovarian cancer (OC) is the most lethal gynecological cancer with a 46% survival rate five years after diagnosis (1). The poor prognosis is related to the fact that most of the patients (75%) get their diagnosis first in the advanced stage of the disease. Partly, this is due to the asymptomatic nature of early-stage OC. Today there are limited diagnostic and imaging tools available to distinguish between cystadenomas and cystadenocarcinomas, which prevents a correct and early diagnosis and subsequent appropriate treatment. The most frequently used marker for OC in blood is the mucin glycoprotein MUC16 (CA125). CA125/MUC16 has high prognostic value for post therapy follow up but has poor sensitivity for diagnosing ovarian carcinomas in the early stages.

The presence of mucin-type glycoproteins in ovarian tumor tissue and cyst fluid have been known for more than 70 years, and they have been shown to carry ABO and Lewis blood group glycan motifs (2). Using proteomics and immunohistochemistry, we and others have shown that in addition to CA125/MUC16 there are other mucins in ovarian tumor cyst fluids, namely MUC5AC, MUC5B, MUC6, and MUC1, although their absolute and relative abundances are unknown (3, 4). Mucin glycosylation changes involving truncation of *O*-glycans are observed in many epithelial cancers, most likely providing cancer cells beneficial properties to spread in the body and to avoid detection by the immune system (5-9). The occurrence and potential use of cancer associated carbohydrate epitopes (Tn, sialyl Tn, T-antigen, sialyl Lewis a and sialyl Lewis x) on CA125/MUC16 and MUC1 in serum and tissue sections from OC patients for differential diagnosis is currently being explored (10-12). Less attention has been given the role of sulfation in OC tissue. Increased levels of the glycosaminoglycan chondroitin sulfate (CS), a major constituent of the extracellular matrix, and increased mRNA levels of CS-associated sulfotransferases, have been observed in OC [reviewed in (13, 14)]. The limited availability of analytical tools such as antibodies or lectins used for detection and quantitation of sulfated *O*- or *N*-glycans have hindered research within this field.

We have previously characterized the glycosylation on mucin-type proteins from benign and malignant mucinous and serous ovarian tumor cyst fluids using a mass spectrometric approach (15), where we observed that the highest level of glycan sulfation was found on proteins from serous cystadenomas. Here, we aimed to further investigate sulfation of Gal-residues on *O*-glycans of IC glycoproteins. The sulfated *O*-glycans collected from ovarian tumor cyst fluids from two patients, one diagnosed with a serous benign adenoma, and one diagnosed with a low-grade serous carcinoma (LGSC), were sequenced using LC-MS^n^ experiments. Using recombinant expression, we tested the specificity of three Gal-sulfotransferase candidates on *O*-glycans, and used serous ovarian tissue microarrays (TMA) probed with antibodies against sulfotransferase candidates, we addressed the question of sulfotransferase regulation during OC development.

## EXPERIMENTAL PROCEDURES

Chemicals were from Merck, Sigma-Aldrich if not otherwise stated.

### Clinical samples

Ovarian tissue samples and cyst fluids were collected after patients signed informed consent. The collection, storage and analysis procedures were approved by the local Ethics Committee at Sahlgrenska University Hospital (Dnr: S348-02 and S445-08) and abide by the Declaration of Helsinki principles. Ovarian tumor cyst fluids used for LC-MS^n^ were purified from two patients as described elsewhere (15) from one patient diagnosed with serous cystadenoma and one with a low-grade serous carcinoma (LGSC) of FIGO stage IIIC (age range of LC-MS^n^ analysed patients was 55-85 with plasma CA125 8-2500 U/ml). Data of the patient cohort for the TMA are summarized in Supplemental Table 4.

### Preparation of desialylated *O*-glycans from ovarian tumor cyst fluids

Acidic glycoproteins from ovarian tumor cyst fluid samples were purified as described elsewhere (15). Briefly, acidic glycoproteins were extracted from ovarian tumor cyst fluids using DEAE anion exchange chromatography followed by ethanol precipitation. Glycoproteins were resuspended in 3.5 M urea, and dot-blotted onto polyvinylidene fluoride (PVDF) membranes and stained with Direct Blue 71. Oligosaccharides were released by reductive β-elimination from the PVDF membrane as described elsewhere (16). The reduced glycans were chemically desialylated using 2% acetic acid for 45 minutes at 95°C.

### Cell culture, expression vectors, transfection and clonal selection, production and purification of PSGL/mIgG2b fusion proteins

To evaluate the specificities of candidate sulfotransferases (CHST1, Gal3ST2 and Gal3ST4) in vitro, plasmids encoding these sulfotransferases were transiently expressed in CHO-K1 cells (ATCC, Manassas, VA, USA) together with plasmids encoding the P-selectin glycoprotein ligand-1/mouse immunoglobulin G2b (PSGL-1/mIgG2b) fusion protein, with or without the plasmid encoding the β1,6-*N*-acetylglucosaminyltransferase 1 (GCNT1) responsible for the synthesis of core 2 *O*-glycans. CHO-K1 cells were cultured in 75-cm^2^ T-flasks (Nunc, Roskilde, Denmark) in Dulbecco’s modified Eagle’s medium (DMEM, Lonza Group Ltd., Basel, Switzerland) supplemented with 10% fetal bovine serum (FBS, Invitrogen AB, Stockholm, Sweden) and were transfected 24 hrs later at a cell confluence of 70-80%. The cells were transfected using the Lipofectamine 2000 Transfection Reagent Kit (Invitrogen). Twenty-four micrograms of plasmid were used for transfection of one 75-cm^2^ flask. Plasmids encoding human galactose-3-O-sulfotransferase 2 and 4 mRNA (pEZ-M67-Gal3ST2 and pEZ-M67-Gal3ST4) were obtained from GeneCopoeia (Rockville, MD, USA). The human keratan sulfate Gal-6-sulfotransferase (CHST1) was amplified by PCR from a human placenta cDNA library using forward (5’-CGCGGG<u>AAGCTT</u>ACCATGCAATGTTCCTGGAAGG-3’) and reverse (5’-CGCG<u>GCGGCCGC</u>TCACGAGAAGGGGCGGAAGTC-3’) primers and was swapped into a CDM 8-based vector carrying the CMV promoter using *Hind* III/*Not* I restriction sites. The plasmids harboring PSGL-1/mIgG2b and GCNT1 were constructed as described previously (17). After an 8-day incubation in 15 ml DMEM with 10% FBS, the supernatant was collected and cells removed by centrifugation at 5020 g for 30 min. The different recombinant PSGL-1/mIgG2b fusion proteins were purified from 15 mL of supernatant by mixing with 30 µL of goat anti-mouse IgG agarose beads in a tube rotated at 4°C for 6 hr. The beads were washed two times with phosphate buffered saline (PBS) and boiled in LDS non-reducing sample buffer (30 µL, Invitrogen AB) before SDS-PAGE electrophoresis and *O*-glycan analysis. In brief, recombinant proteins were separated on 3-8% NuPAGE gels (Invitrogen AB) and electrophoretically blotted onto PVDF membrane (Immobilon P membranes, Millipore, Bellerica, MA) using a semi-dry method. *O*-glycans were released from PVDF membrane stripes using reductive β-elimination as described previously (16, 18).

### LC-MS of *O*-glycans and data assignments

For LC-MS, the glycans were separated on porous graphitized carbon columns (PGC, 5 µm particles, Hypercarb, ThermoFisher Scientific, Waltham, MA, USA) prepared in-house, and with 10 cm length and 250 µm inner diameter. The gradient was 0-40% acetonitrile containing 10 mM ammonium bicarbonate over 60 min, kept at a flow rate of 5-10 μL/min. Mass spectrometric data was collected at low resolution in the negative ion mode using a Thermo Scientific LTQ ion trap mass spectrometer (San Jose, CA).

MS (collision induced dissociation) analyses of *O*-glycans were performed by repeated injections of the same sample and analyzed with data-dependent MS^n^ experiments. The glycans were detected as deprotonated ([M-nH]^n-^) ions. For the LC runs involving MS^2^ experiments, MS^2^ was performed on the six most abundant ions in every full scan or performed throughout the run on a predetermined ion of interest. For LC runs involving MS^3^ and MS^4^ analyses (ovarian tumor cyst fluid glycans), data-dependent MS experiments were performed where each target ion was continuously selected for fragmentation throughout the whole run (summarized in Supplemental Table 3). Constant parameters were electrospray voltage 3.5 kV, capillary voltage of -6V, and capillary temperature of 300°C. Full MS scans were collected at *m/z* 380-2000 with an isolation width of *m/z* 2, and normalized collision energy of 35%.

MS spectra were interpreted manually, and the assignment of sequences and annotation of fragment ions were confirmed using freely available software ‘GlycoWorkbench’(19). Reference spectra for sulfated core 1 sequences were collected from PSGL-1/mIgG2b purified from supernatants of transient CHO-K1 transfectants expressing sulfotransferases Gal3ST4, CHST1 and PSGL1/mIgG2b without co-transfection with GCNT1 (see section on recombinant proteins). The following assumptions were made based upon known features of mucin-type *O*-glycosylation: Hex residues are Gal, HexNAc are GlcNAc, reducing end sugar is GalNAc-ol.

### Tissue Micro Array (TMA) and scoring

Sulfotransferase protein expression levels in serous ovarian tissues were evaluated using TMA with tissue from 323 ovarian tumor specimens (Supplemental Table S4). The construction and scanning of the TMA is described elsewhere (20). The paraffin sections were deparaffinated in xylene and then rehydrated. Tris-base pH 9 was used as unmasking solution. The sections were counterstained with Hematoxylin.

The tissue sections were incubated with polyclonal antibodies produced in rabbit: Anti-CHST1 (ARP45495_T100, Aviva Systems Biology), anti-Gal3ST2 or anti-Gal3ST4 (HPA071809 and HPA038137, Prestige Antibodies supported by Human Protein Atlas, Sigma-Aldrich). The antibody dilutions were for CHST1, 1:100; for Gal3ST2, 1:20; and for Gal3ST4, 1:50 in PBS. They were incubated with the tissue for 90 min. Antibody binding was measured using UltraVision Quanto Detection System AP (Thermo Scientific) according to the protocol.

A semiquantitative method ‘quick score’ (QS) for manual evaluation of tissue staining was applied. Proportion of stained tissue was graded 0-4, based upon percentage of stained tissue within the following intervals: 0 = 0%, 1 = 1-25%, 2 = 26-50%, 3 = 51-75% and 4 = 76-100%. Intensity of the stain was graded 0-3 (negative, weak, moderate, and strong). The scores for proportion and intensity were summed, generating a maximum score value of 7. Every tissue section was scored blindly by pathologist (CM) in duplicates or triplicates.

### Experimental Design and Statistical Rationale

Chemical desialylation of oligosaccharides from cyst fluids from two patients with benign and malignant epithelial ovarial tumors was performed once due to limited availability. These samples were injected 17 and 14 times (benign and malignant, respectively) each for the targeted MS^n^ experiments outlined in Supplemental Table 3. The OC MS data presented in this paper is primarily presented as descriptive. For the study of the specificity of sulfotransferases on recombinantly produced PSGL1/ mIgG2b, the data presented here were generated from one transfection with sulfotransferase Gal3ST2 or two transfections performed with CHST1 and Gal3ST4. Glycosylation from each transfection was analyzed with LC-MS once. TMA sulfotransferase staining and intensity scoring was performed once on the full sample set due to limited availability and validated on a subset (n=28) of serous OC tissue on a separate TMA. Human colon and placenta tissue slides were used as positive controls.

## RESULTS

### Analyses of desialylated sulfated oligosaccharides from serous ovarian tumor cyst fluids

Oligosaccharides were released by reductive β-elimination from enriched acidic proteins from cyst fluids from two patients, one patient diagnosed with benign tumor, and one diagnosed with malignant LGSC. The oligosaccharides were desialylated to aid MS identification of sulfate location. The *O*-glycans were subjected to analyses with LC-MS^n^ using porous graphitized carbon chromatography and negative ion mode. Base peak chromatograms are displayed in Figure 1A and B, where major peaks are assigned. The LC-MS profiles confirmed that sulfated glycans were abundant in these samples. The degree of sialylation before removal of these residues is compiled in Supplemental Table S2, and also described within a previous study (15). Major sulfated glycan precursor ions were identified, followed by repeated analyses of the samples using data-dependent MS^n^ experiments to ensure sufficient number of MS^2^, MS^3^ and MS^4^ spectra on the low abundant sulfated compounds. Fifty-four sulfated glycans consisting of 2-10 monosaccharide residues with 1-4 sulfate residues were detected, many as isomeric structures. The general structural theme of identified oligosaccharides is shown in Figure 1C, and the compiled list of deduced sequences is found in Supplemental Table S1.

**Figure 1.**
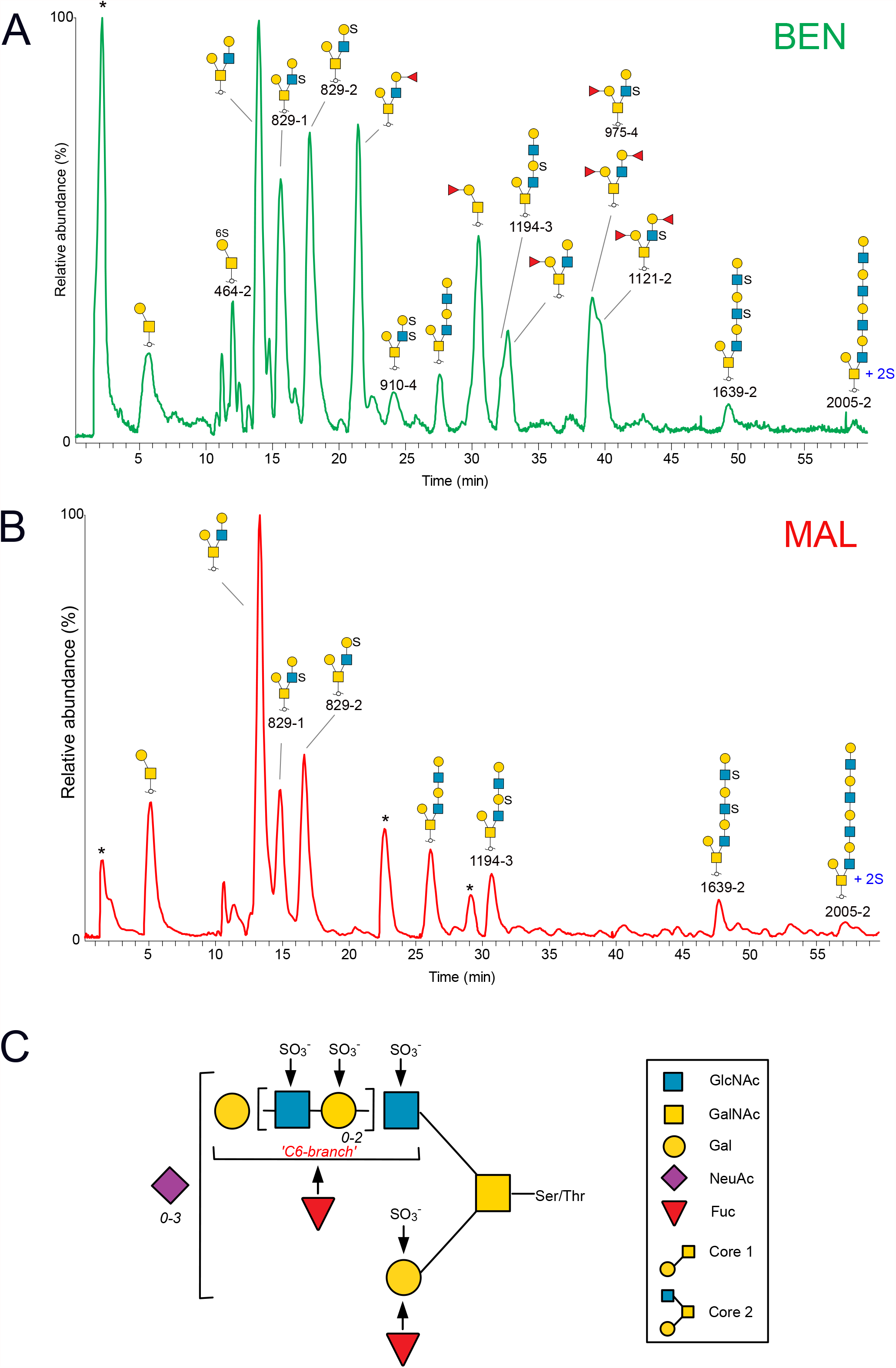
LC-MS base peak chromatograms of desialylated sulfated glycoforms, purified from serous ovarian tumor cyst fluids from patients with serous benign tumor (A, ‘BEN’) and malignant tumor (B, ‘MAL’), respectively. Major glycans are annotated using SNFG nomenclature. Sulfated glycans are annotated with name in red font and are listed in Supplemental Table 1. Contaminants are labelled with *. In C, general structural theme of sulfated *O*-glycans from serous ovarian tumor cyst fluids is displayed. The GalNAc residue is linked to serine or threonine in the protein. Residues marked with arrows indicate identified sites for fucosylation or sulfation. Numbers in italics indicate that Gal-GlcNAc (highlighted in brackets) and NeuAc residues can occur 0, 1, 2 and 3 times in the molecule. Symbol key is according to the SNFG nomenclature for glycans (36).

### Core 1 glycans from serous ovarian tumor cyst fluids carry sulfate linked to hydroxyl groups at C-3 and C-6 of Gal

Two sulfated core 1 type glycans were detected in the serous ovarian tumor cyst fluid samples, two monosulfated disaccharides (HSO_3_ -Galβ1-3GalNAcol) detected as [M-H]^-^ ions at *m/z* 464 labelled ‘464-1’ and ‘464-2’ (Supplemental Table S1). The extracted ion chromatograms from the two patients are inserted in Figure 2, highlighting that the component retention times and the relative abundances were similar in both samples. The corresponding MS^2^ spectra were identical in the two samples, supporting that the same glycans were present. MS^2^ spectra of ‘464-1’ and ‘464-2’ from the serous benign tumor cyst fluids are shown in Figure 2A and B. Both spectra contained an intense C type fragment ions at *m/z* 241^-^, supportive of sulfate predominantly linked to the Gal residue, and the late eluting sequence ‘464-1’ displayed low intense Gal-cross ring fragment ions at *m/z* 199^-^ (^0,2^A) shown previously to be formed when sulfate is linked via C-6 to Gal (21). The two glycoforms were deduced as ‘HSO_3_-3Galβ1-3GalNAcol’ (464-1’) and ‘HSO_3_-6Galβ1-3GalNAcol (‘464-1’) by comparison to MS^2^ spectra of reference compounds, obtained from recombinantly produced PSGL1/mIgG2b fusion protein transfected into CHO-cells together with sulfotransferases CHST1 or Gal3ST4. (Supplemental Figure 1).

**Figure 2.**
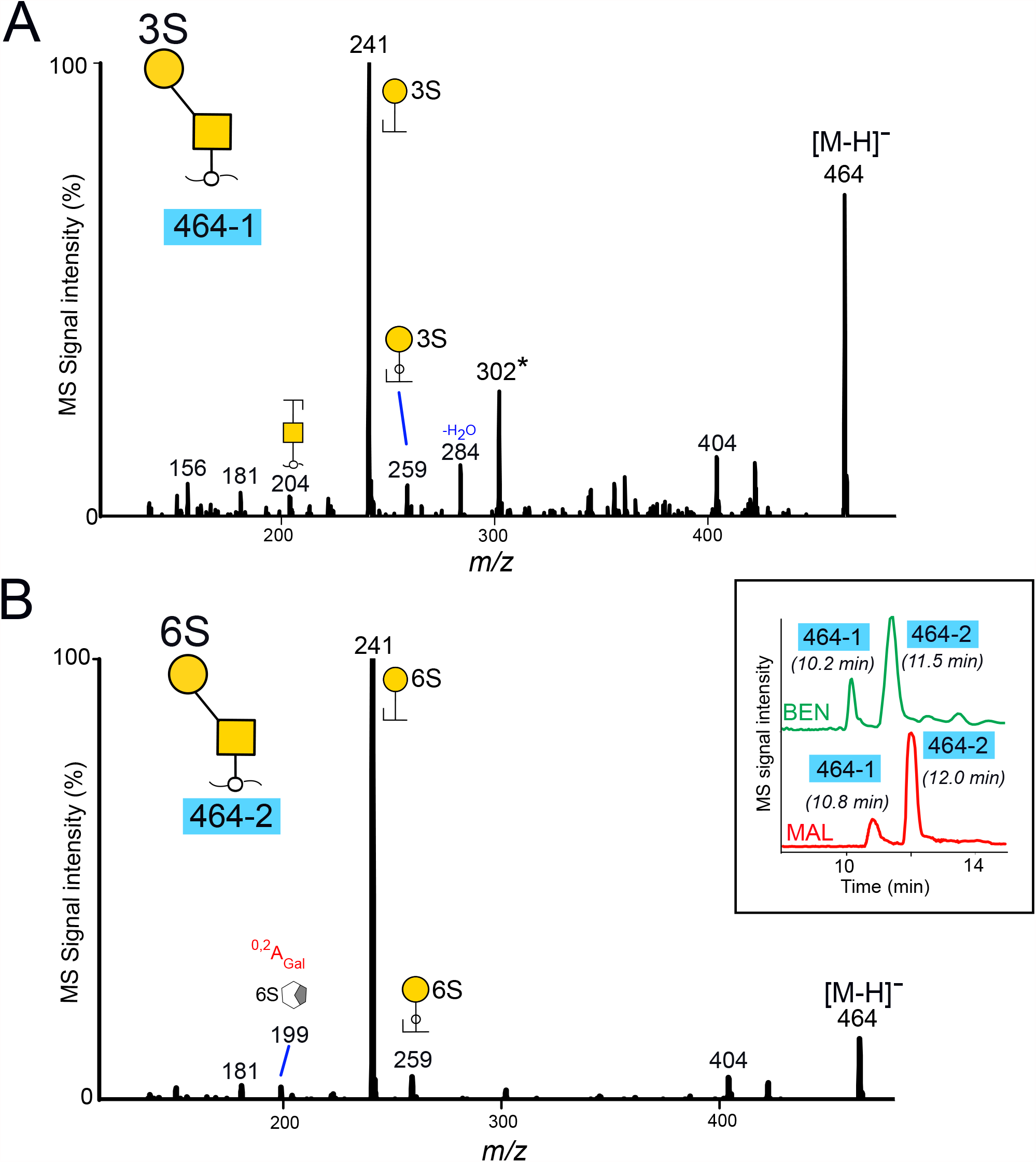
MS^2^ spectra of sulfated core 1 disaccharides at *m/z* 464 ([M-H]^-^ ion) labelled as ‘464-1’(A) and ‘464-2’ (B), purified from cyst fluids from OC patients with benign (‘BEN’) and malignant (‘MAL’) tumors. The extracted ion chromatograms from the full scans are inserted. For key of symbols, see Figure 1. * The ions at *m/z* 302 in panel A corresponds to a fragment ion consisting of sulfate linked to the core reducing sugar GalNAcol, and may origin from minor amounts of a coeluting glycoform.

Evidence for a third, lower abundant glycoform co-eluting with ‘464-1’ (Figure 2A) and present in the samples from both the benign and the malignant tumor cyst fluids, where a fragment ion at *m/z* 302 indicative of sulfate linked to the core GalNAcol, was observed (22).

### Core 2 glycans from serous benign and malignant tumor cyst fluids have sulfate linked to both GlcNAc and Gal residues

The majority of the desialylated glycans detected from cyst fluids were assigned as core 2 type with a single Gal residue linked via C-3 to GalNAcol, and an extended C6 branch (Figure 1C). Examples of two short, sulfated saccharides are displayed in Figure 3. Extracted ion chromatograms of two abundant tetrasaccharides detected at *m/z* 829 ([M-H]^-^ precursor ions) from the two patients are inserted in the figure. The two components which were labelled ‘829-1’ and ‘829-2’ had similar retention times, relative abundances, and the same MS spectra in both samples. Both glycans were interpreted as core 2 sequences, where the most intense fragment ions corresponded to loss of a Hex residue (*m/z* 667^-^). The ions at *m/z* 444^-^ in both spectra were indicative of a branch consisting of Hex, HexNAc and sulfate. The ions at *m/z* 505^-^ and 282^-^ in 829-1 (Figure 3A) supported a core 2 sequence with sulfate linked to HexNAc, and thus interpreted as Gal-3(Gal-(HSO_3_-)GlcNAc-6)GalNAcol. Component ‘829-2’ (Figure 3B) contained fragment ions at *m/z* 241^-^, indicative of sulfate linked to Hex, and the sequence assumed to be Gal-3(HSO_3_-Gal-GlcNAc-6)GalNAcol. The spectra did not reveal fragment ions which enabled assignment of C-3 or C-6 linked sulfate. Additional identified structures also showed sulfation either on Gal and GlcNAc residues or both (Supplemental Table S1).

**Figure 3.**
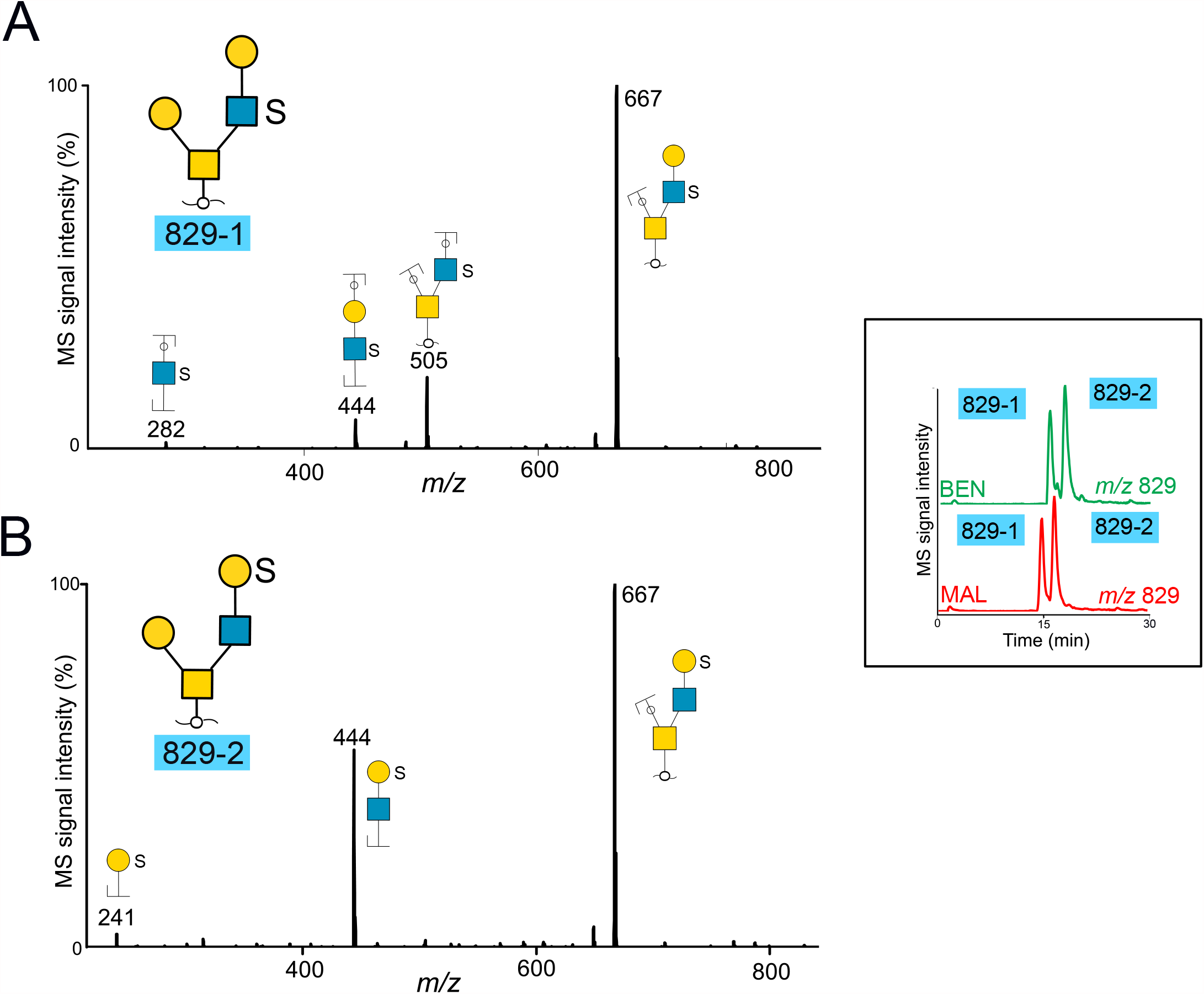
MS2 spectra of desialylated sulfated core 2 tetrasaccharides detected at *m/z* 829 and labelled ‘829-1’ (A) and ‘829-2’ (B), purified from cyst fluids from two patients with benign (‘BEN’) and malignant (‘MAL’) tumors. The parent ions were detected as [M-H]^-^ ions. The extracted ion chromatograms are inserted. For key of symbols, see Figure 1.

### Data dependent MS^n^ experiments revealed doubly, triply and tetrasulfated glycans with a single Hex linked on the core 2 C-3 branch and extended LacNAc chain linked to the C-6 branch

A general structural theme of the core 2 type sulfated oligosaccharides was found in cyst fluids from the two patients with benign and malignant tumors, with the glycan chain extension restricted to the C-6 arm, and consisting of up to four Hex-HexNAc units (Figure 1C and Supplemental Table S1). Increasing number of sulfates caused the glycans to bind strongly to the porous graphitized carbon (PGC) column. The largest glycans that were detected here consisted of five Hex, five HexNAc and two sulfate residues (‘2005-1, -2 and - 3’, Supplemental Table S1). These eluted within a minute before the gradient ended, precluding detection of larger glycans eluting later.

In many cases the sulfate residues could be assigned to inner Gal or GlcNAc residues in the glycan. This was supported by the detection of an abundant fragment ion in MS^2^ spectra generated from the loss of a terminal non-sulfated Hex, followed by a second MS^3^ collision experiment of this ion, displaying the loss of a second, non-sulfated Hex. Most of these compositions were identified to contain sialic acid before desialylation (Supplementary Table 2), suggesting that terminal Hex residues are sialylated. An example of the type of structure found is illustrated in Figure 4. Figure 4A shows the base peak chromatogram of the sulfated glycoforms detected at *m/z* 819^2-^/1639^-^, originating in malignant cyst fluid proteins. The monosaccharide composition corresponded to oligosaccharides consisting of eight residues and two sulfate groups. The deduced sequences in Figure 4A are listed in Supplemental Table S1. All were found to be of core 2 type, with a single non-sulfated Hex linked to the reducing HexNAcol, and a second extended disulfated triLacNAc chain carrying sulfate in different positions causing the observed structural variety. The complexity is illustrated by at least five detected chromatographic peaks in Figure 4A, labelled ‘1639-1’ to ‘1639-5’. The most abundant component (‘1639-2’
s) was found to be sulfated on the two outer HexNAc residues in the triLacNAc unit. Of these ‘1639’ isomers, the ‘1639-2’ was the dominating isomer detected in the sample purified from the patient with a benign tumor (Supplemental Table 1). Figure 4B displays MS^2^ spectra of one of the minor components labelled ‘1639-3’ only present in the malignant sample. The major fragment ions at *m*/i 738.1^2-^ originated from loss of a terminal Hex residue. It is surrounded by singly and doubly charged fragment ions from the extended branch (B/C-ions) on the C-6 arm or containing the reducing sugar (Y/Z-ions). The presence of intense C-ions at *m/z* 626.6^2-^ confirmed the presence of the C6 branch, consisting of six monosaccharide residues and the two sulfates. Figure 4C shows the MS^3^ spectra for component ‘1639-3’ collected by the data-dependent MS experiment covering the transition ‘819.3^2-^ → 738.1^2-^’ ([M-Hex]^2-^). The most abundant fragment ions at *m/z* 657.2^2-^, were indicative of loss of a second, non-sulfated, terminal Hex residue from the precursor molecule. The detection of sulfated fragment ions at *m/z* 241^-^, 282^-^ and 261.5^2-^, supported that those sulfates were linked to a Hex and a HexNAc, respectively, and that these saccharide residues had to be adjacent to each other. Assignment of the disulfated Hex-HexNAc unit within the linear chain was aided by detection of doubly charged B-ions at *m/z* 525.2^2-^, which indicated that this unit had to be part of the five outer monosaccharide residues of the extended C-6 branch. Detection of Y-ions at *m/z* 829^-^ and 870^-^ in the MS^2^ and MS^3^ spectra confirmed that one sulfate was present on the first Hex closest to the reducing sugar in the linear C6-branch. Figure 4D displays MS^4^ spectra covering the transition ‘819.3^2-^ → 738.1^2-^ → 657.2^2-^’ ([M-2Hex]^2-^), thus fragmenting the precursor oligosaccharide at 657.2^2-^ which has already lost two Hex residues. These spectra confirmed our interpretation of the sequence, since all the detected fragment ions could be deduced from the remaining residues.

**Figure 4.**
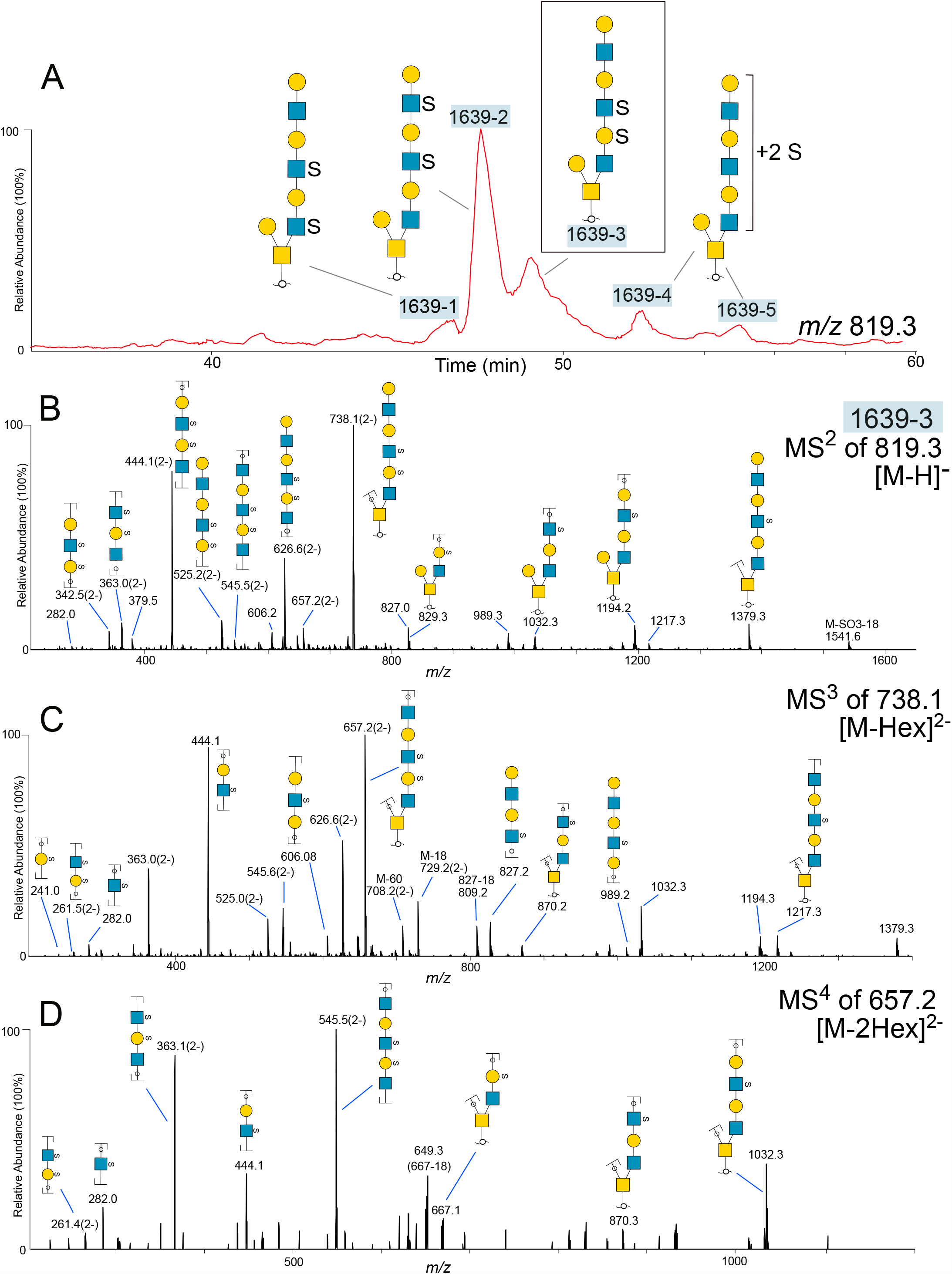
(A) Extracted ion chromatograms of desialylated disulfated oligosaccharides detected at *m/z* 819, purified from cyst fluid from a patient with a malignant (‘MAL’) tumor. The parent ions were detected as [M-2H]^2-^ ions. Glycan sequences are interpreted from MS^2^, MS^3^ and MS^4^ experiments. (B) MS^2^ spectra of the glycan labelled “1639-3”. Fragment ions are annotated with proposed compositions. For some fragment ions, more than one composition may be possible. (C) MS^3^ spectra of the ions at *m/z* 738.1^2-^ from the MS^2^ experiment shown in (B). (D) MS^4^ spectra of the ions at *m/z* 657.2^2-^ from the MS^3^ experiment shown in (C). For key of symbols, see Figure 1.

Assignment of sulfate residues along the extended C6-chain of the sulfated glycans was aided by the detection of abundant doubly charged fragment ions carrying two sulfate groups, such as *m/z* 261^2-^ consisting of two sulfates and a Hex-HexNAc unit (B-ions), and *m/z* 373^2-^, doubly sulfated Z-ions consisting of Hex-HexNAc and the reducing sugar HexNAcol. A confirmation of these diagnostic ions was provided in MS^2^, MS^3^ and MS^4^ experiments of the disulfated tetrasaccharide labelled ‘910-4’ (Supplemental Table S1). This glycan was interpreted as Gal-3(HSO_3_ -Gal-(HSO_3_ -)GlcNAc-6)GalNAcol, and the MS^n^ spectra are shown in Supplemental Fig. S2. The doubly charged fragment ions at *m/z* 373^2-^ were the most abundant ions in the MS^2^ spectra, and was further collided in an MS^3^ experiment, revealing fragment ions at *m/z* 261^2-^, supporting that the sulfate groups were present on Hex-HexNAc. B-ions at *m/z* 241^-^ confirmed that one sulfate was linked to a Hex, and Y/Y-ions at *m/z* 505^-^ that the second sulfate was linked to a HexNAc. MS^4^ of *m/z* 261^2-^ as a precursor revealed fragment ions confirming both the saccharide and sulfate constituents of this fragment.

### The pattern of desialylated, sulfated O-glycans from serous benign and malignant ovarian tumor cyst fluids revealed no major differences regarding non-fucosylated sulfated structures

Figure 5 shows the extracted MS base peak chromatograms of six different glycan compositions, consisting of between 4 and 8 monosaccharide residues and 2-3 sulfate residues. The deduced compositions are shown above and are also found listed in Supplemental Table S1. Overall, the same number of peaks and approximate relative abundance are observed in both samples, with one exception. One of the two glycoforms of the trisulfated tetrasaccharides shown in panel 989 is not detected in the cyst fluid sample from the patient with a malignant tumor. However, in all, these glycan profiles do not point towards any major differences regarding relative abundances of sulfated glycans from the benign and malignant cyst fluid samples, although minor differences are observed which could be associated with altered sulfotransferase activity.

**Figure 5.**
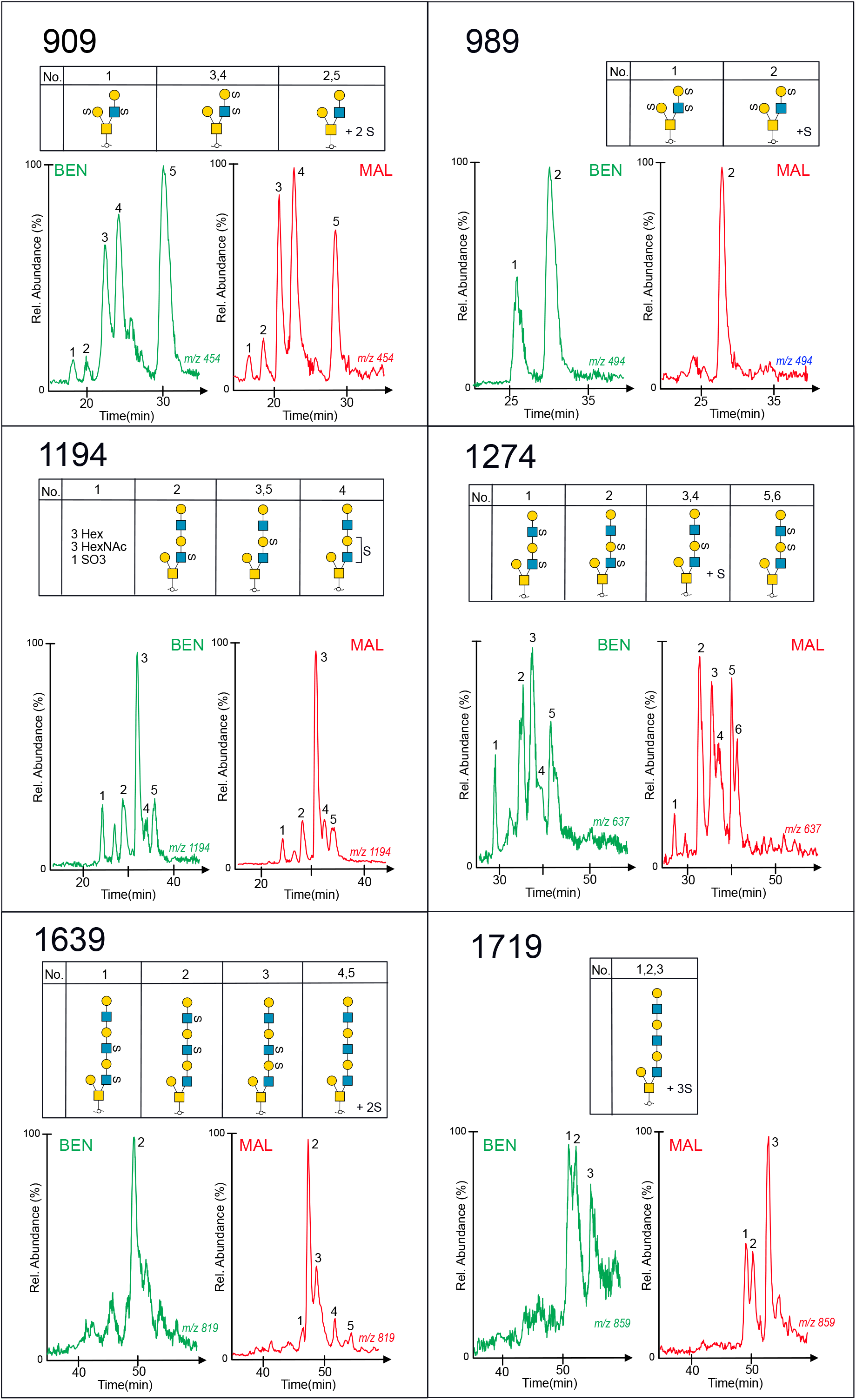
Extracted ion chromatograms of desialylated sulfated glycoforms, purified from serous cyst fluids from OC patients with benign (labelled in red) and malignant (labelled in blue) tumors. Glycan sequences were interpreted from MS^2^, MS^3^ and MS^4^ experiments. Glycan panel names (909, 989, 1194, 1274, 1639 and 1719) corresponds to their mass in the deprotonated form and are listed in Supplemental Table S1. For key of symbols, see Figure 1.

### Fucose-containing sulfated glycans provide evidence for sulfated Lewis b/y and blood group H sequences in serous benign cyst fluid but only Lewis a/x sequences in malignant cyst fluid

Desialylation of serous cyst *O*-glycans also enabled structural characterization of some of the low abundant fucose-and sulfate-containing glycans present on serous cyst fluid proteins. Figure 6 displays the base peak chromatograms of both a non-sulfated and a sulfated fucosylated pentasaccharide (panels 895 and 975) from benign and malignant cyst fluids. Sequence elucidation confirmed that the detected structures were both of blood group H (Fucα1-2Gal-) and Lewis (Galβ1-3/4(Fucα1-4/3)GlcNAc-) type. The base peak chromatogram (Panel 1121) of the difucosylated sulfated hexasaccharide revealed two glycoforms detected in the benign but not in the malignant cyst sample. The glycoform labelled ‘1121-1’ was of a Lewis b/y type (Fucα1-2Galβ1-3/4(Fucα1-4/3)GlcNAc-), and the corresponding MS^2^ spectra of the precursor ions at *m/z* 1121 ([M-H]^1-^) showed multiple diagnostic and intense fragment ions used for sequence identification. The B-ions at *m/z* 736.1^-^ indicated the presence of a branch containing two Fuc, sulfate, Hex, and HexNAc, and the Y-ion at *m/z* 959^-^, formed by loss of a non-substituted, terminal Hex residue. The fragment ions at *m/z* 505^-^ confirmed that sulfate was present on the HexNAc adjacent to the reducing sugar, and the ions at *m/z* 428^-^ was diagnostic of a HexNAc substituted with both sulfate and Fuc. The sequence was thus deduced as equivalent to a core 2 Lewis b or Lewis y sequence: Galβ1-3(Fucα1-2Galβ1-3/4(Fucα1-4/3)(HSO3-)GlcNAcβ1-6)GalNAc.

**Figure 6.**
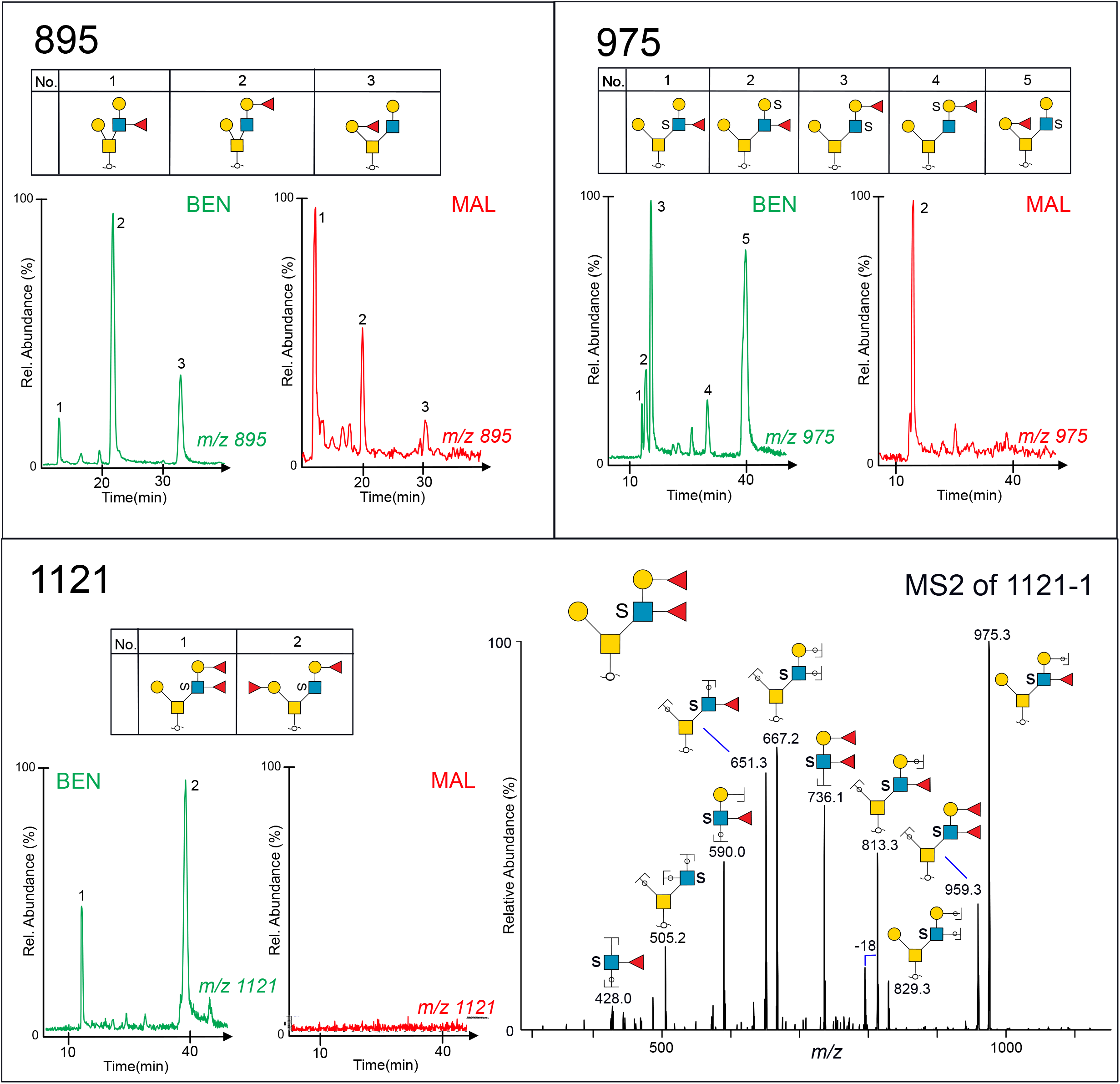
Extracted ion chromatograms of desialylated fucose- and sulfate-containing glycoforms, purified from cyst fluids from benign (labelled in red) and malignant ovarian tumors (labelled in blue). Glycan sequences were interpreted from MS^2^ experiments. Glycan panel names (895, 975, 1121) corresponds to their mass in the deprotonated form and are listed in Supplemental Table 1. MS^2^ of a sulfated Lewis b or Lewis y type glycan “1121-1” detected as *m/z* 1121 ([M-H]^-^ precursor ion) is displayed in the lower panel. For key of symbols, see Figure 1.

### Sulfotransferases CHST1, Gal3ST2, and Gal3ST4 add sulfate to C-3 and C-6 of different Gal residues in core 2 O-glycans on PSGL-1/mIgG2b fusion proteins

Our results highlighted that sulfate linked to Gal residues both within and at the terminal non-reducing end of the *O*-glycan branches is a significant feature of ovarian glycoproteins. In order to test if some of the identified sulfotransferases described in the literature were able to use mucin core 1 and/or 2 *O*-glycans as targets, PSGL-1/mIgG2b was co-expressed with the core 2 glycosyltransferase (GCNT1) and the Gal sulfotransferases Gal3ST2, Gal3ST4 or CHST1 in CHO cells. The three sulfotransferases have been shown to add sulfate to C-3 (Gal3ST2 and Gal3ST4) and C-6 (CHST1) of Gal. We have previously published the *O*-glycosylation of PSGL-1/mIgG2b expressed in CHO cells together with the core 2 GCNT1 but without sulfotransferases, and the glycans were found to be non-sialylated, mono- and disialylated core 1 and core 2 glycans consisting of 3-6 monosaccharide residues (17). PSGL-1/mIgG2b from the three transfections were purified, the *O*-glycans released using reductive β-elimination, followed by analyses with LC-MS and MS^2^ experiments. Interpretations are compiled in Supplemental Figure 3. The extracted ion chromatograms and MS^2^ spectra of the core 2 tetrasaccharide with the sulfate added to the sequence Galβ1-3(Galβ1-4GlcNAcβ1-6)GalNAcol detected as *m/z* 829^-^ ([M-H]^-^ ions) are shown in Figure 7. All three sulfotransferases generated only one glycoform product each. All three MS^2^ spectra revealed fragments ion at *m/z* 241^-^, diagnostic of sulfate linked to a Hex residue, but the detected glycoforms were interpreted differently, as shown. The spectra of the glycoform produced by the Gal3ST2 sulfotransferase contained additional fragment ions at *m/z* 444^-^, indicative of sulfate linked to Hex in the LacNAc unit on the C6 branch. The MS^2^ spectra for the other two sulfotransferase glycoforms contained fragment ions at *m/z* 464^-^, indicative of sulfate attached to Gal linked C-3 to the reducing sugar GalNAc. The two glycoforms generated by Gal3ST4 and CHST1, which differed only by C-3 or C-6 linked sulfate, had identical retention times and identical MS^2^ spectra. In addition to the 829 glycoforms, a few sialylated sulfated glycans were detected. The CHST1 sulfotransferase added two sulfates to a core 2 glycan detected at *m/z* 599^2-^, the MS^2^ spectra was interpreted NeuAcα2-3(HSO_3_-6)Galβ1-3(SO_3_-6Galβ1-4GlcNAcβ1-6GalNAcol. This indicated that CHST1 could add sulfate to Gal also to the LacNAc C6-linked chain.

**Figure 7.**
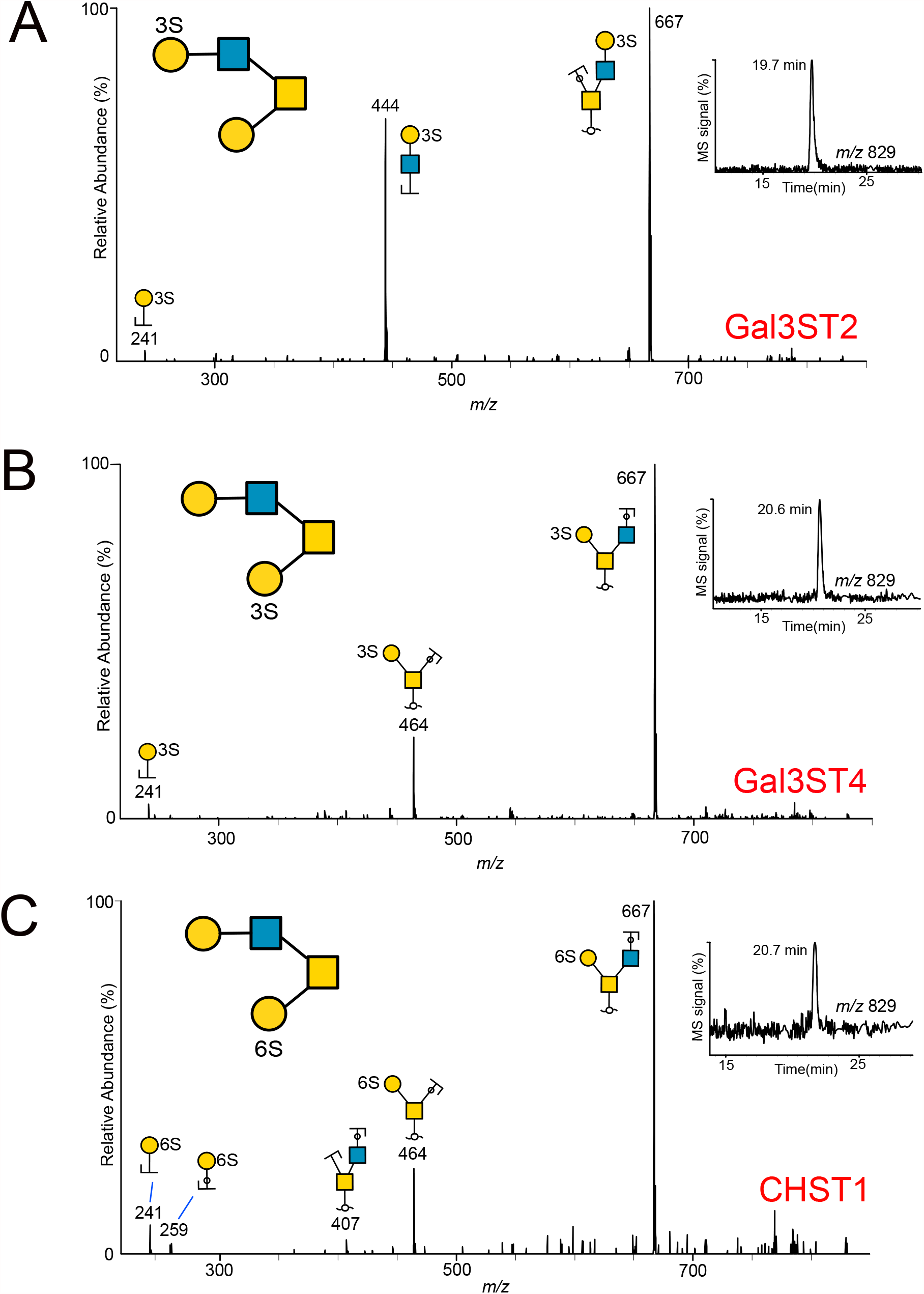
MS^2^ spectra of sulfated core 2 *O*-glycans collected after fragmentation of the precursor ion at *m/z* 829 ([M-H]^-^) from recombinantly produced PSGL-1/mIgG2b protein purified from CHO cells. PSGL-1 was co-transfected with core 2 transferase (GCNT1) and sulfotransferases Gal3ST2 (A), Gal3ST4 (B) or CHST1 (C). For key of symbols, see Figure 1.

In summary, these experiments confirm that Gal3ST2, Gal3ST4 and CHST1 all add sulfate to Gal residues of the core 2 Galβ1-3(Galβ1-3GlcNAcβ1-6)GalNAc precursor present on the recombinantly produced PSGL1/mIgG2b in CHO cells, although with different fine specificities.

### Sulfotransferase Gal3ST2 is decreased in tissue from serous ovarian adenocarcinomas compared to serous ovarian adenomas

To further investigate the pathological relevance of the three Gal sulfotransferases in ovarian tissue, we screened patients (n=323) using immunohistochemistry in TMA with specific antibodies targeting each of the three sulfotransferases. Representative immunohistochemical staining of tissue from two patients with malignant cancer and one benign patient are displayed in Figure 8A. The three graphs in Figure 8B depict the groups consisting of selected patients with benign, borderline, and malignant epithelial ovarian tumors. The picture also includes a numerical score of a combined assessment of intensity of staining and % positive cells (quick score QS). The results showed that all the three sulfotransferases are present in the three classes of ovarian tumors. No significant differences in the scoring were detected for CHST1 and Gal3ST4, whereas Gal3ST2 displayed a significant decrease (P<0.0001) with a higher median QS value in benign compared to malignant tumor tissue.

**Figure 8.**
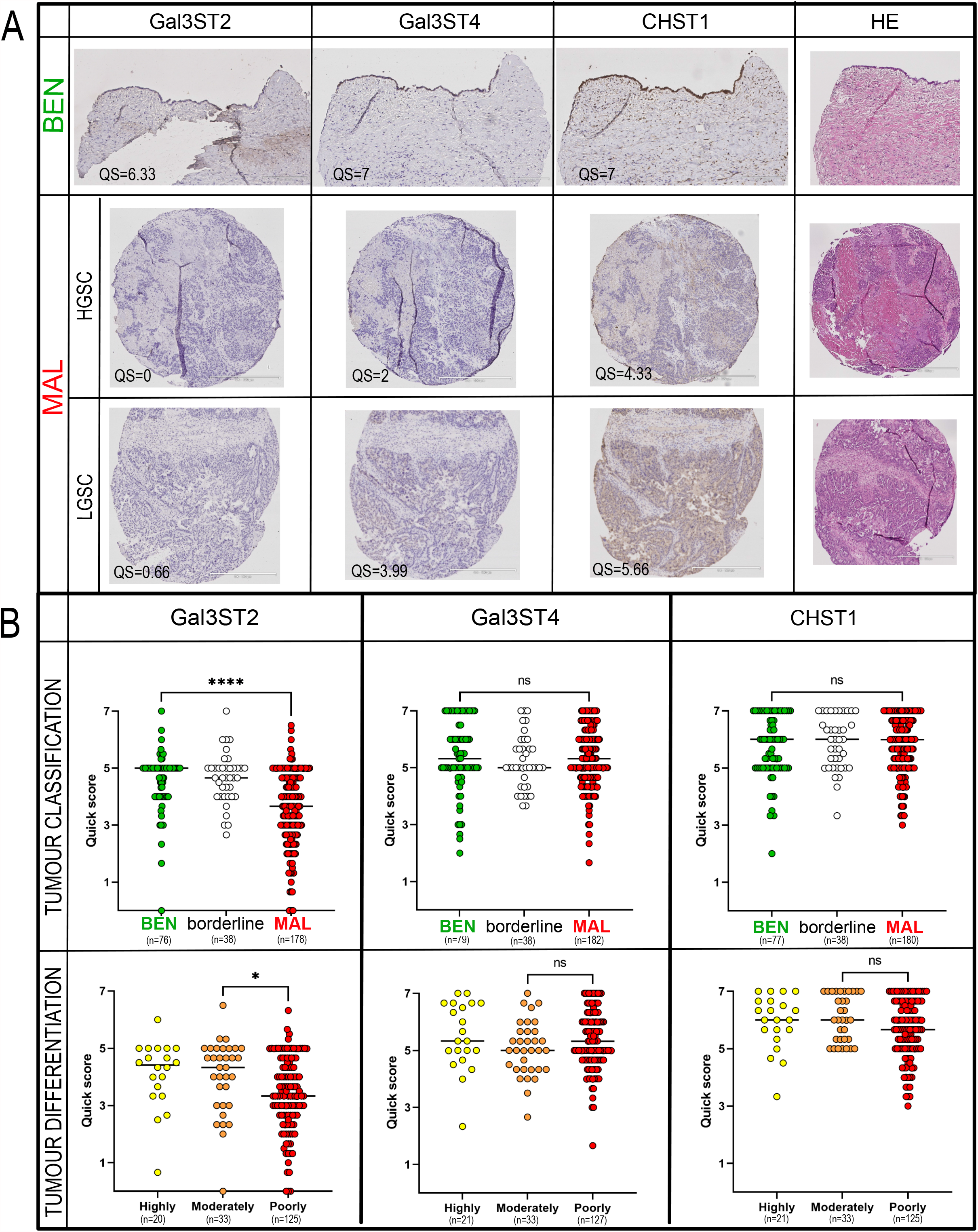
Protein expression of sulfotransferases Gal3ST2, Gal3ST4 and CHST1 in benign and malignant serous ovarian tumor cells analyzed using immunohistochemistry and tissue micro arrays. (A) Representative antibody staining of tissue from patient diagnosed with serous cystadenoma, patient with low-grade serous carcinoma (LGSC, stage IIC, type 1), and patient with high grade serous carcinoma (HGSC, stage IIIC, type 2). Immuno-positive staining is observed as brown coloring of tissue. HE = hematoxylin-eosin. Scale bars represent 200 µm (benign tissue sections) or 500 µm (malignant tissue sections). (B) Tissue micro array data was based upon the screening of tissue from 323 patients. The clinicopathological data is compiled in Supplemental Table S4. The number of patients in each group as defined by malignancy grade (benign (‘BEN’), borderline, malignant (‘MAL’) or tumor differentiation (‘highly’, ‘moderately’ or ‘poorly’) according to WHO 2008 classification is shown in the figure (37). Quick score (QS) scoring (0-7) was based upon manual evaluation and the sum of the proportion and intensity of staining (see materials and methods section). Tissue sections were analysed in duplicates or triplicates. The bar in the graphs indicate the median value. Significant differences were calculated by Mann-Whitney test.

## DISCUSSION

In our previous study, we concluded that fucosylated and sulfated *O*-glycans were potential biomarkers on ovarian serous cyst fluid proteins (15). However, the blood group associated enzymes FUT2 and FUT3 in secretions are not expressed in all individuals. In for example caucasians, approximately 80% express a functional FUT2, and approximately 90% express a functional FUT3. Blood group ABO fucosylation was found to be closely related to the mucinous subtype of OC. Here, we have focused on structural assignment of sulfated glycans from the serous type. The *O*-glycans were desialylated and we performed data-dependent MS^2^, MS^3^ and MS^4^ experiments, which allowed structural elucidation. Desialylation was performed since sialic acid prevents detailed characterization of sulfation using MS in two ways. First, they give rise to more structural glycoforms which lowers the relative abundance of each component. Second, CID-spectra of larger sialylated glycans are often not very informative, since most collision energy is spent on dissociation of the NeuAc glycosidic bonds.

In our previous study, we reported an overall estimated decrease in the abundance of sulfated glycans on serous cyst fluid proteins from patients diagnosed with malignant forms of OC (15). In the current study, we have performed a detailed structural assignment of sulfated *O*-glycans derived from cyst fluid proteins of two patients, one classified as benign, the second as malignant, grade ‘LGSC’. The latter was chosen to include a malignant cancer, where we could detect significant amount of sulfation, while in the more aggressive ‘HGSC’ tumor, their lower level of sulfation would have meant that sulfated glycans would be more difficult to characterize.

Our experiments revealed that ovarian glycoproteins carried sulfated core 1 and 2 *O*-glycans consisting of up to ten residues, with 1-4 sulfates linked via hydroxyl groups to *N*-acetylhexosamine residues or C-3 and C-6 to Gal in repeated LacNAc units on the C-6 branch. The presence of disulfated LacNAc units on glycoproteins from ovarian malignant tissue has previously been described by Shibata el al. (23). Based upon the LC-MS chromatograms of the desialylated sulfated oligosaccharides, no major qualitative or quantitative differences between the two samples could be observed.

Chromatographic profiles of short monosulfated glycans such as the core 1 sulfated T-antigens (HSO_3_ +Galβ1-3GalNAc) (Figure 2) and sulfated core 2 glycans (HSO_3_ +Galβ1-3(Gal-GlcNAcβ1-6)GalNAc) (Figure 3) were similar between the two samples analysed. This was also valid for the larger, sulfated glycans shown in Figure 5, with one exception. However, this exception could not be simply explained relating to an activity of a single sulfotransferase, and it remains as an unexplained observation that have to be adresserd on a larger cohort of sampoles. However, as an overall conclusion, MS data indicate that it is the same bulk of sulfated glycans that are present in both the benign and malignant cyst fluid samples. Our previous results (15) that pointed towards a decreased level sulfation in malignant ovarian tumors suggests in this report to be mainly due to an overall decrease of sulfation rather than caused by altered activity of a single sulfotransferase, at least not a Gal-S transferase. However, in this report with the aim to thoroughly characterize the nature of sulfation in OC using only two patient samples, extended conclusion of the sulfoglycome in serous OC cannot be drawn.

At current, it is unclear to which proteins ovarian glycans are attached to, and if the sulfation manifest differently on different protein cores. Prime protein candidates for sulfation are the mucin type proteins revealed by immunohistochemistry and proteomics, such as CA125/MUC16, MUC5AC, MUC5B, MUC6, and MUC1 (3, 4).

We also investigated the relative abundance of sulfotransferases in ovarian tissue between different patient stages. There are 37 human Golgi membrane bound sulfotransferases reported (24), many have primarily been studied due to their involvement in the sulfation of proteoglycans and/or glycolipids, whereas less is known about their precursor substrate specificity and activity towards glycoproteins. Sulfated glycan motifs on glycoproteins are involved in interactions between leukocytes and the endothelium, such as L-selectin mediated re-circulaton of lymphocytes to lymph nodes or leukocyte extravasation at sites of inflammation, and as ligands for sialic acid binding immunoglobulin like receptors (Siglecs) which participate in the discrimination between ‘self’ and ‘nonself’ (25, 26). For mucins, which are commonly found as gel-forming or transmembrane proteins at mucosal surfaces, sulfated glycans have been proposed to be involved in the interaction and protection against microbiota (27, 28).

We detected sulfate linked to both Gal and GlcNAc in these samples, and showed that sulfate could be linked both C-3 and C-6 to Gal. Since sulfation on Gal on glycoproteins is less well studied, we chose to explore three sulfotransferases: CHST1, Gal3ST2 and Gal3ST4. Extended mucin type *O*-glycans are commonly composed of repeated poly *N*-acetyllactosamine units [polyLacNAc; (−4GlcNAcβ1-3Galβ1-)_*n*_], which restricts Gal-3 sulfation only to terminally positioned residues, whereas Gal-6 sulfation occurs both on internal and terminal Gal. CHST1 (synonyms KSGal6ST, C6ST) was cloned 1997 (29). Its activity was confirmed and characterized by *in vitro* experiments observing the incorporation of ^35^S at the C6-position of Gal in keratan sulfate, but at a higher rate, to non-glycosaminoglycans on fetuin and to C-6 of Gal in sialyl *N*-acetyllactosamine oligosaccharides (NeuAcα2-3Galβ1-4GlcNAcβ1-) (30). It has been explored for its involvement in the biosynthesis of L-selectin ligands on high endothelial venules (31). During characterization of a monoclonal antibody HMOCC-1 against the ovarian clear cell cancer cell line RMG1, CHST1 was found to play a regulatory role in the formation of the sulfated glycan HMOCC-1 epitope (23). We showed here that CHST1 is extensively expressed in most OC tissues (Figure 8). We confirmed its specificity acting on core 2 glycans on PSGL-1/IgG expressed in CHO cells, showing that the enzyme may add sulfate to C-6 of both Gal residues of the core 2 tetrasaccharide NeuAcα2-3Galβ1-3(Galβ1-GlcNAcβ1-6)GalNAc.

There are four Gal-3-sulfo transferases described of which three (Gal3ST2, Gal3ST3 and Gal3ST4) are found to act on glycoproteins, and all are found in ovarian tissue. Gal3ST2 (GP3ST) was shown to add sulfate to C-3 of Gal in synthetic or purified oligosaccharides, preferentially on type 1 (-Galβ1-3GlcNAc-) and type 2 (-Galβ1-4GlcNAc-) chains and to a lesser extent on the T-antigen (Galβ1-3GalNAc) (32). This is in line with our findings, where Gal3ST2 preferentially adds sulfate to the Gal in the LacNAc unit linked C-6 to GalNAc in the core 2 tetrasaccharide (Figure 7). Analogous, we have previously reported core and branch dependent specificity for *O*-linked glycans for the α1,3 fucosyltransferase family (33) using the CHO-cell expression approach. Gal3ST4 was initially cloned and characterized 2001 by Seko *et al*. (34). He reported the incorporation of sulfate to C-3 of Gal linked to GalNAc in the core 2 saccharide Galβ1-3(GlcNAcβ1-6)GalNAc, which is concordant to our results. Gal3ST3 was not included in the current study, but is reported to act on core 2 glycans (35) and nextprot.org reports RNA levels present in ovarian tissue. The involvement of this enzyme in OC requires further investigation.

Our TMA study showed that protein expression levels of sulfotransferases Gal3ST4 and CHST1 remained the same in serous ovarian tissue from all tumor stages and degree of differentiation, whereas Gal3ST2 levels were decreased in malignant tissue. A decrease in sulfotransferase levels is in line with our previous findings where we used a mass spectrometric approach to semi-quantify sulfated glycans from eight patients (15). However, our MS data indicates that there is no major change in the sulfated glycoforms in the samples from different patient groups, and it is likely that the overall sulfation decrease as previously indicated (15) must be related to more factors than Gal3ST2, which only can sulfate terminal Gal. There are additional sulfotransferase candidates which may add sulfate to C-3 or C-6 in Gal of glycoproteins and are reported to be expressed in ovarian tissue (Gal3ST3 and CHST3). This may influence the overall sulfation, and further investigation of the role of sulfotransferases adding sulfate to C-6 of GlcNAc residues will provide complementary information about the role and regulation of sulfation in serous OC.

## Data Availability

Raw MS data and Glycoworkbench files which includes fragment ion assignment for ovarian cancer cyst fluid O-glycan analyses are available on glycopost ( https://glycopost.glycosmos.org/) using the project ID: GPST000192. Structures with available MIRAGE information are available at Unicarb-DR (https://unicarb-dr.glycosmos.org/references/453).
Raw MS data for analyses of O-glycans from PSGL1/mIg2b co transfected with sulfotransferases Gal3T2, Gal3ST4 and CHST1 and with or without core 2 transferase GCNT1 are available at glycopost using the project IDS: GPST000198 and GPST000199, respectively. Structures with available MIRAGE information are available at Unicarb-DR, reference numbers 456 (core2+CHST1), 457 (core2+Gal3ST2), 458 (core2+Gal3ST4), 459(core1+Gal3ST4) and 460(core1+CHST1).

https://glycopost.glycosmos.org/

https://unicarb-dr.glycosmos.org/references/453

https://unicarb-dr.glycosmos.org/references/456

https://unicarb-dr.glycosmos.org/references/457

https://unicarb-dr.glycosmos.org/references/458

https://unicarb-dr.glycosmos.org/references/459

## Data Availability

https://glycopost.glycosmos.org/

https://unicarb-dr.glycosmos.org/references/453

https://unicarb-dr.glycosmos.org/references/456

https://unicarb-dr.glycosmos.org/references/457

https://unicarb-dr.glycosmos.org/references/458

https://unicarb-dr.glycosmos.org/references/459

## Data Availability

https://glycopost.glycosmos.org/

https://unicarb-dr.glycosmos.org/references/453

https://unicarb-dr.glycosmos.org/references/456

https://unicarb-dr.glycosmos.org/references/457

https://unicarb-dr.glycosmos.org/references/458

https://unicarb-dr.glycosmos.org/references/459

## AUTHOR CONTRIBUTIONS

VV purified and analyzed ovarian glycans, KAT interpreted the spectra. JL, CJ and JH designed and made the PSGL constructs. CJ purified the rePSGL and analyzed the glycans. NGK, VV and KS generated the funding for the research. NGK and KS developed the research strategy and directed and managed the project. KS is responsible for sample collection. KA and CM designed the TMA. BW constructed the TMA and performed the immunohistochemistry. CM performed the pathological assessment and evaluated the TMA staining of the TMA. KAT performed the statistical analysis of the TMA. KAT, NGK and KS together wrote the manuscript, all authors discussed the results, commented and approved the final manuscript.

## DATA AVAILABILITY

Raw MS data and Glycoworkbench files which includes fragment ion assignment for OC cyst fluid *O*-glycan analyses are available on glycopost (https://glycopost.glycosmos.org/) using the project ID: GPST000192. Structures with available MIRAGE information are available at Unicarb-DR (https://unicarb-dr.glycosmos.org/references/453). Raw MS data for analyses of *O*-glycans from PSGL1/mIg2b co transfected with sulfotransferases Gal3T2, Gal3ST4 and CHST1 and with or without core 2 transferase GCNT1 are available at glycopost using the project IDS: GPST000198 and GPST000199, respectively. Structures with available MIRAGE information are available at Unicarb-DR, reference numbers 456 (core2+CHST1), 457 (core2+Gal3ST2), 458 (core2+Gal3ST4), 459(core1+Gal3ST4) and 460(core1+CHST1).

## SUPPLEMENTAL DATA

This article contains supplemental data. Supplemental Figures S1 to S3 and Tables S1 to S4.

## Abbreviations

OC: ovarian cancer;
HGSC: high grade serous carcinoma,
LGSC: low-grade serous carcinoma;
Tn antigen: GalNAc-á-Threonine/Serine;
T antigen: Galâ1-3GalNAc-Threonine/Serine;
sLea: sialyl Lewis a;
sLex: sialyl Lewis x;
Hex: hexose;
HexNAc: Nacetylhexosamine;
LC-MS: liquid chromatography-mass spectrometry;
CHO cells: chinese hamster ovary cells;
TMA: tissue micro array

