## Supplementary data for "Sulfation of *O*-glycans on mucin-type proteins from serous ovarian epithelial tumors"

### Supplemental Material

**Figure S1**. Extracted LC-MS base peak chromatograms and $MS^2$ spectra of components '$SO_3$-3Galβ1-3GalNAcol' (A) and '$SO_3$-6Galβ1-3GalNAcol' (B) detected at *m/z* 464 ($[M\text{-}H]^-$ parent ion).

**Figure S2**. MS spectra of the oligosaccharide detected at *m/z* 454 and labelled "910-4" (Table S1) purified from cyst fluid from patient with a malignant tumor.

**Figure S3**. *O*-glycans detected on recombinant PSGL-1/mIgG2b produced in CHO cells, cotransfected with core 2 transferase (GCNT1), and sulfotransferases Gal3ST2, Gal3ST4 or CHST1.

**Table S1.** Sulfated desialylated *O*-glycans from ovarian serous cyst fluid proteins from two patients diagnosed with cystadenoma ('Benign') or papillary cystadenocarcinoma ('Malignant').

**Table S2.** Sulfated *O*-glycans with from ovarian serous cyst fluids from two patients diagnosed with cystadenoma ('Benign') or papillary cystadenocarcinoma ('Malignant').

**Table S3.** Compilation of data-dependent MS experiments performed during separate LC-MS runs, used in this project to characterize desialylated sulfated reduced *O*-glycans from ovarian serous cyst fluid proteins from two individuals with cystadenoma ('Benign') or papillary cystadenocarcinoma ('Malignant').

**Table S4.** Clinicopathological characteristics of serous tumors evaluated for immunohistological stainings with sulfotransferases Gal3ST2, Gal3ST4 or CHST1.

**Figure S1**. Extracted LC-MS base peak chromatograms and MS$^2$ spectra of components 'SO$_3$-3Galβ1-3GalNAcol' (A) and 'SO$_3$-6Galβ1-3GalNAcol' (B) detected at *m/z* 464 ([M-H]$^-$ parent ion). The glycans were purified from recombinantly produced 'PSGL-1' protein in CHO cells, co-transfected with sulfotransferases galactose-3-O-sulfotransferase 4 ('Gal3ST4') which adds sulfate via a hydroxyl group C-3 to Gal (A), or with carbohydrate sulfotransferase 1 ('CHST1') which adds sulfate to C-6 to Gal (B).

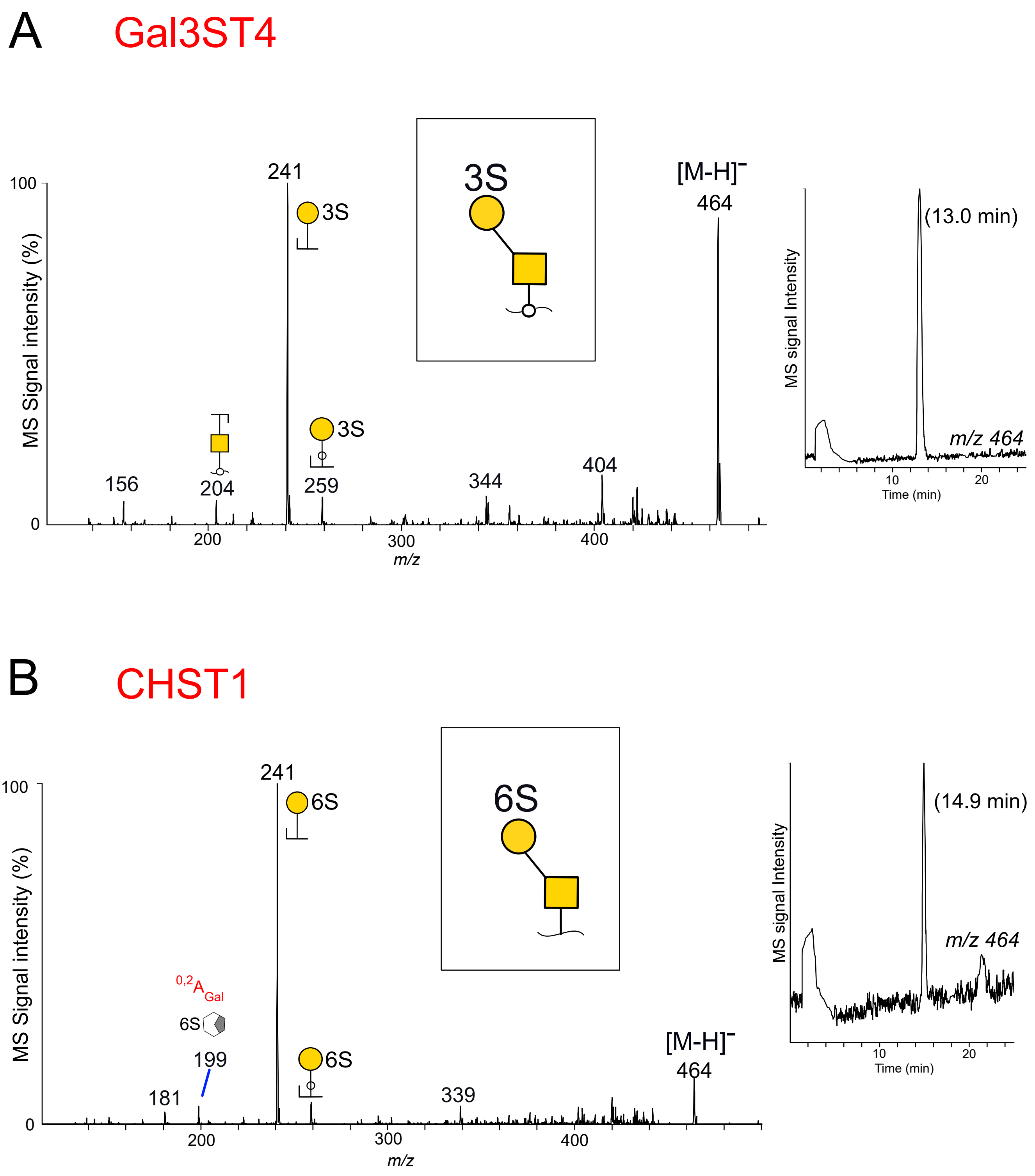

**Figure S2**. (A) MS2 spectra of the oligosaccharide detected at *m/z* 454 and labelled “910-4” (Table S1) purified from cyst fluid from patient with a malignant tumor. The extracted LC-MS base peak chromatogram from the full scan is inserted. Fragment ions are annotated with proposed compositions. For some fragment ions, more than one composition may be possible. (B) $MS^3$ spectra of the ion at *m/z* $373.1^{2-}$ from the $MS^2$ experiment shown in (A). (C) $MS^4$ spectra of the doubly charged ion at *m/z* $261^{2-}$ from the $MS^3$ experiment shown in (B). For key of symbols, see Figure 1.

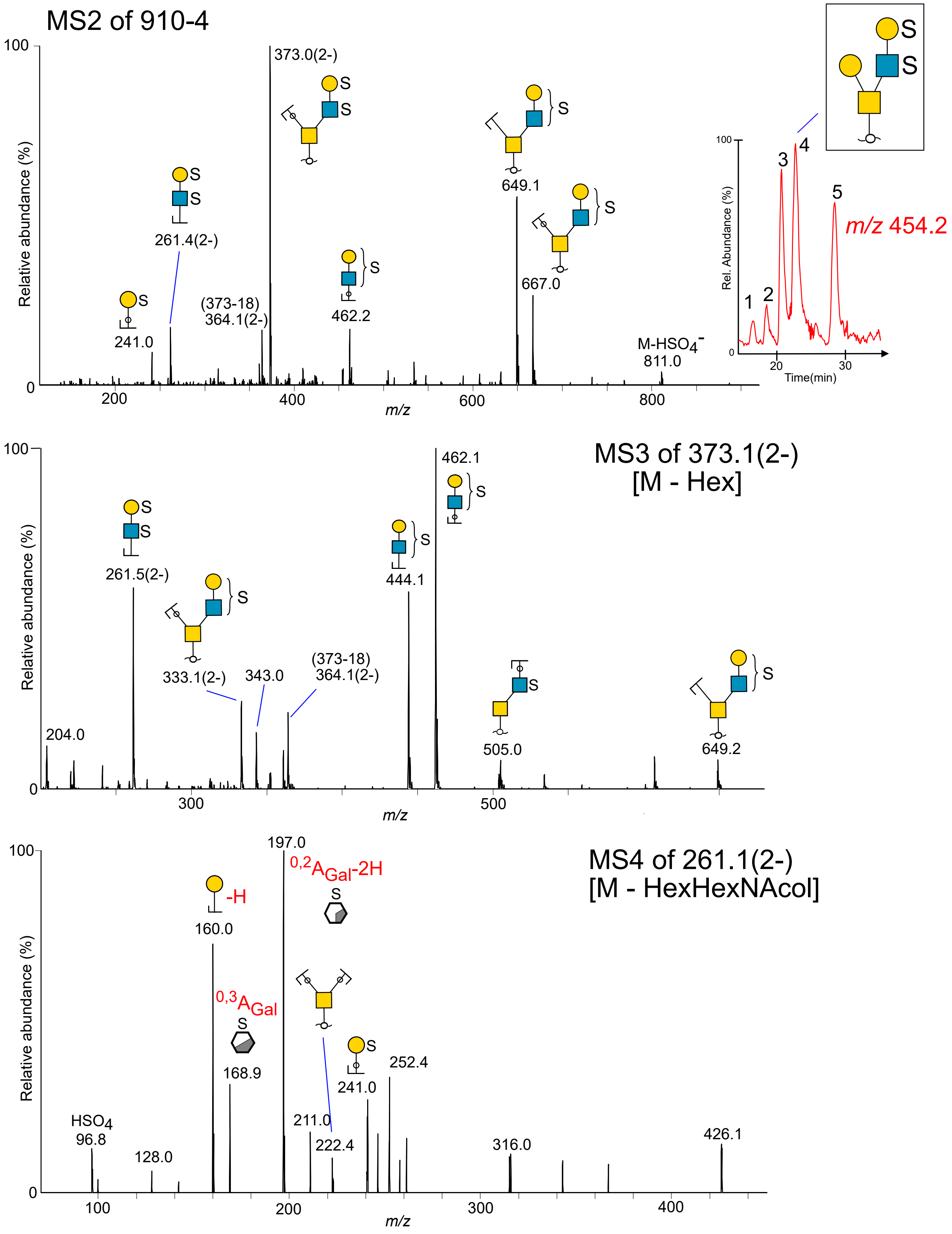

**Figure S3**. *O*-glycans detected on recombinant PSGL-1/mIgG2b produced in CHO cells, cotransfected with core 2 transferase (GCNT1), and sulfotransferases Gal3ST2, Gal3ST4 or CHST1. The glycans were analyzed with LC-MS and $MS^2$ experiments. The spectra were interpreted manually and compared to reference spectra, as described in Experimental Procedures. Relative amounts of different glycans are given in percentage (%) of the total sum of integrated peak areas from one (Gal3ST2) or two (CHST1 and Gal3ST4) separate transfections. [1]O-glycans detected on PSGL-1/mIgG2b + GCNT1 and their relative amounts are from our previous paper and were listed here as reference (17). Symbol key is found in Figure 1.

| *m/z* *[M-xH]$^{x-}$* | PSGL-1/mIgG2b + GCNT1[1] | + GAL3ST2 | + GAL3ST4 | + CHST1 |
|---|---|---|---|---|
| 675 | 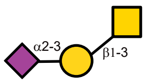  2% | 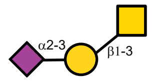  8% | 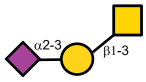  11% | 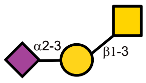  10% |
| 749 | 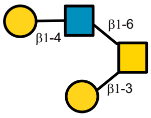  1% | | 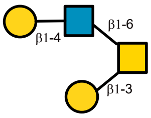  6% | 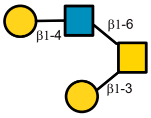  6% |
| 878 | 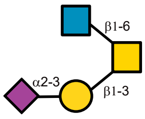  2% | 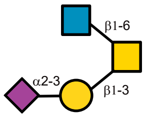  6% | 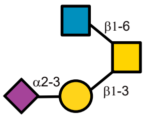  8% | 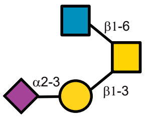  5% |
| 1040 | 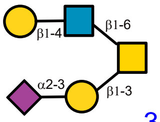  39% | | 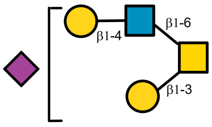  40% | 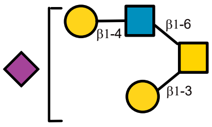  24% |
| 1331 | 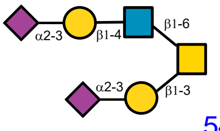  54% | | 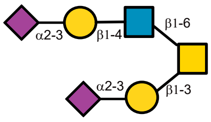  30% | 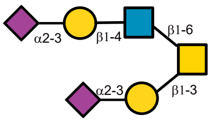  16% |
| 829 | | 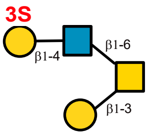  6% | 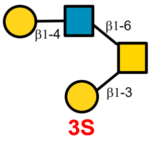  5% | 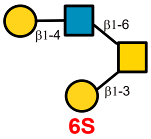  3% |
| 1120 | | 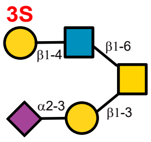  79% | | 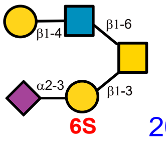  20% |
| 599 | | | | 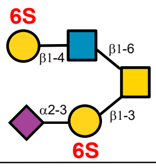  17% |
